# Genome Wide Analysis Across Alzheimer’s Disease Endophenotypes: Main Effects and Stage Specific Interactions

**DOI:** 10.1101/2021.08.13.21261887

**Authors:** Tanner Y. Jacobson, Kwangsik Nho, Shannon L. Risacher, Sujuan Gao, Li Shen, Tatiana Foroud, Andrew J. Saykin, for the Alzheimer’s Disease Neuroimaging Initiative

**Affiliations:** Indiana Alzheimer’s Disease Research Center, Indianapolis, IN 46202, USA; Department of Medical and Molecular Genetics, Indiana University School of Medicine, Indianapolis, IN 46202, USA; Department of Radiology and Imaging Sciences, Indiana University School of Medicine, Indianapolis, IN 46202, USA; School of Informatics and Computing, Indiana University School of Medicine, Indianapolis, IN 46202, USA; Department of Biostatistics, Indiana University School of Medicine, Indianapolis, IN 46202, USA; Department of Biostatistics, Epidemiology and Informatics, The Perelman School of Medicine, Philadelphia, PA 19104 USA

**Keywords:** Genetics, GWAS, endophenotype, *APOE*, Genetic Interaction, Cerebrospinal Fluid Biomarkers, Magnetic Resonance Imaging, Amyloid-PET, FDG-PET

## Abstract

**Introduction:** Genetic association analysis of key Alzheimer’s disease (AD) endophenotypes may provide insight into molecular mechanisms and genetic contributions.

**Methods:** Major AD endophenotypes based on the A/T/N (Amyloid-β, Tau, and Neurodegeneration) biomarkers and cognitive performance were selected from Alzheimer’s Disease Neuroimaging Initiative (ADNI) in up to 1,565 subjects. Genome-wide association analysis of quantitative phenotypes was performed using a main SNP effect and a SNP by Diagnosis interaction (SNPxDX) model to identify stage specific genetic effects.

**Results:** Sixteen novel or replicated loci were identified in the main effect model, with six (*SRSF10, MAPT, XKR3, KIAA1671, ZNF826P*, and *LOC100507506*) meeting study significance thresholds with the A/T/N biomarkers. The SNPxDX model identified three study significant genetic loci (*BACH2, EP300, PACRG-AS1*) associated with a neuroprotective effect in later AD stage endophenotypes.

**Discussion:** An endophenotype approach identified novel genetic associations and new insights into the associations that may otherwise be missed using conventional case-control models.

## 1 BACKGROUND

The complex nature of Alzheimer’s disease (AD) has instigated multiple studies that have identified genetic risk factors and potential contributors to the disease, but the underlying drivers for the disease are not yet completely understood. The known genetic risk factors do not fully explain the genetic heritability of AD. As such, new approaches are needed to further identify the underlying genetic players in AD.

One such approach is the use of biomarker endophenotypes to increase statistical detection power and to provide valuable insight into the molecular mechanisms underlying AD [1]. An ideal phenotype lies downstream of the genetics and upstream of the “observable” phenotype, in this case cognitive changes in AD. They provide quantifiable measures of the pathology of the disease for genetic association.

The Alzheimer’s Disease Neuroimaging Initiative (ADNI) provides a wide variety of biomarkers highly associated with AD, and several studies have utilized this dataset for genetic analysis [2]. The completion of ADNI phases, ADNI1, GO, and ADNI2, provide an opportunity to evaluate multiple endophenotypes together in a shared dataset, allowing a comprehensive analysis of genetic associations with AD endophenotypes.

In this study, two models were used to evaluate genetic associations with AD endophenotypes: A main effect model of overall genetic associations with baseline measures of endophenotypes and a SNP by Diagnosis (SNPxDX) interaction model of genetic associations within diagnostic stages of AD. The main effect model follows a traditional linear regression approach applied to the individual biomarkers, and has the power to identify overall genetic effects on endophenotypic measures across all subjects and diagnostic classification groups.

The SNPxDX model analyzes the relation between genetic variation and endophenotype in the context of stage specific clinical diagnostic classification: cognitively normal (CN), early mild cognitive impairment (EMCI), late mild cognitive impairment (LMCI), and AD. Progression along the stages of the clinical syndrome of AD is not uniform across patients, with evidence that genetic factors influence this disease heterogeneity [3, 4]. Past work in the ADNI genetic core has suggested an interaction with specific known AD genes and diagnostic group, but a robust genome wide analysis across multiple phenotypes has not been performed [5]. The main effect model is well suited for identifying genetic effects that influence endophenotype change across all subjects and diagnostic groups, providing a contrast between individuals that are cognitively normal and those that are on the AD spectrum of diagnosis. With the heterogenous nature of AD and pathophysiological changes that occur over the course of the disease genetic effects that are stronger or only occur in specific stages of disease progression may be reduced in the analysis. Introducing an interaction term between genetic marker and the diagnostic classification may provide more power to identify stage specific genetic effects.

There is increasing evidence that late onset AD is largely determined by multiple, small effect and low penetrance genetic factors [6, 7]. Identifying genetic effects that influence AD disease risk is important for developing an understanding of the disease. Applying alternative models to identify genetic risk factors that influence the disease in a complex manner contributes to our overall understanding of the disease and helps pave the path toward therapeutic approaches. Furthermore, identifying how and where, through an endophenotype approach, as well as when, through stage specific interactions, a genetic variant is affecting disease risk enables more focused research in understanding disease mechanisms.

## 2 METHODS

### 2.1 Study Participants

Participants from the Alzheimer’s Disease Neuroimaging Initiative Phase 1 (ADNI-1) and its subsequent extensions (ADNI-GO/2) [8] were included in this study. Further information about these studies, participant enrollment, protocols, and other information can be found at www.adni-info.org. Written informed consent was obtained from each participant, and all protocols were approved by each site’s Institutional Review Board.

### 2.2 Genotyping and Imputation

Whole blood samples from the ADNI participants were genotyped on the Illumina Human 610-Quad BeadChip, the Illumina HumanOmniExpress Beadchip, or the Illumina Omni 2.5M platform (Illumina, Inc., San Diego, CA). After sample and SNP standard quality control procedures of GWAS data [9], genotype imputation and calling was performed over each data set separately using the Haplotype Reference Consortium Panel r1.1. *APOE* genotype for rs429358 and rs7412, described as *APOE* ε2/ε3/ε4 status, were genotyped separately as described previously [5]. To avoid population stratification confounding, non-Hispanic Caucasian ADNI participants (N=1565) were selected for this analysis by genetic clustering using HapMap 3 genotype data and multidimensional scaling (MDS) analysis.

### 2.3 Selected Phenotypes

Biomarkers were selected based on previous studies for association with AD pathology [10]. Baseline measures of 17 phenotypes were selected to represent the key markers of AD represented by A/T/N and C: Amyloid-β (A), Tau (T), Neurodegeneration (N) [11], and an additional Cognitive Performance (C) category. Amyloid-β (A) measures are represented by 1 meta region of interest (ROI) for [18F]Florbetapir amyloid PET and CSF amyloid-β 1-42 peptide (Aβ1-42), Tau (T) measures by CSF total tau (t-tau) and phosphorylated tau (p-tau), Neurodegeneration (N) by MRI atrophy measures (8 ROIs) and FDG PET (3 ROIs), and Cognitive Performance (C) as composite scores developed by Crane et al. [12] for memory (MEM) and executive function (EF). MRI and FDG ROIs were selected based on previous studies of AD pathology and progression and selected to cross-sectionally represent the disease across stages [13, 14]. **Table 1** presents the full list of phenotypes and sample sizes.

**Table 1:** List of selected endophenotypes, their N, and covariates applied during regression analysis indicated by •. * Analysis performed with and without *APOE* covariate. *†* Values were pre-adjusted for MRI-Field strength where applicable.

| Selected Endophenotype | N | Genetic PC1 | Genetic PC2 | APOE E2/E3/E4 Allele* | Age | Sex | Education | Intracranial Volume | MRI Field Strength† |
| --- | --- | --- | --- | --- | --- | --- | --- | --- | --- |
| Baseline Memory Composite Score | 1565 | • | • | • | • | • | • |  |  |
| Baseline Executive Function Composite Score | 1565 | • | • | • | • | • | • |  |  |
| Baseline bilateral mean hippocampus volume | 1555 | • | • | • | • | • | • | • | • |
| Baseline bilateral mean entorhinal cortex thickness | 1555 | • | • | • | • | • | • | • | • |
| Baseline bilateral mean frontal lobe thickness | 1555 | • | • | • | • | • | • | • | • |
| Baseline bilateral mean cingulate thickness | 1555 | • | • | • | • | • | • | • | • |
| Baseline bilateral mean parietal lobe thickness | 1555 | • | • | • | • | • | • | • | • |
| Baseline bilateral mean temp. lobe thickness | 1555 | • | • | • | • | • | • | • | • |
| Baseline bilateral mean medial temp. lobe thickness | 1555 | • | • | • | • | • | • | • | • |
| Baseline bilateral mean lateral temp. lobe thickness | 1555 | • | • | • | • | • | • | • | • |
| Baseline Mean FDG PET SUVR in Angular Gyrus | 1158 | • | • | • | • | • |  |  |  |
| Baseline Mean FDG PET SUVR in Cingulate | 1158 | • | • | • | • | • |  |  |  |
| Baseline Mean FDG PET SUVR in Bilateral Mean Temp. Lobe | 1158 | • | • | • | • | • |  |  |  |
| Baseline [18F]Florbetapir amyloid PET | 791 | • | • | • | • | • |  |  |  |
| Baseline Roche CSF amyloid-β 1-42 peptide | 981 | • | • | • | • | • | | | |
| Baseline Roche CSF Total Tau | 1103 | • | • | • | • | • |  |  |  |
| Baseline Roche CSF Phosphorylated Tau | 1103 | • | • | • | • | • |  |  |  |

### 2.4 Genetic Association Analysis

The main effect genome wide association analysis was performed separately for each phenotype in PLINK v1.9 [15]. Association was evaluated across 5,406,480 genotyped and imputed variants for each phenotype. All phenotypes were adjusted for age, sex, and the first two principal components of the genetic population by inclusion in the linear model. MRI and cognitive measures were additionally adjusted for education, and MRI measures were adjusted for intracranial volume.

The analysis was performed with and without the *APOE* e2/e3/e4 genotype as a covariate to account for the effects of the *APOE* allele on genetic effects. To fully account for an *APOE* genotypic effect, the *APOE* genotype was coded as dummy variables indicating 1 = e2e2, 2 = e2e3, 3 = e3e3, 4 = e2e4, 5 = e3e4, 6 = e4e4.

MRI field strength was identified as an additional covariate for MRI phenotypes; however, MRI field strength was directly tied to ADNI phase with ADNI Phase 1 participants and ADNI 2/GO participants receiving 1.5 Tesla and 3 Tesla MRI, respectively. No significant effect from the ADNI phase was identified in non-MRI measures. Due to different proportions of diagnostic groups within ADNI 1 and 2/GO, a significant over correcting effect was seen in some individuals due to these proportional differences. To address this issue, a regression analysis was run using only individuals identified as cognitively normal, with the MRI phenotype as the dependent variable and age, sex, education, ICV, and MRI field strength as covariates. The resulting Beta coefficient of the MRI Field Strength was then used to adjust the MRI phenotype across all subjects using the following formula:

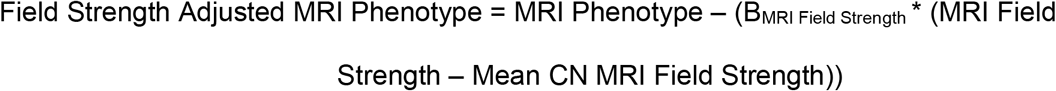

The Field Strength Adjusted MRI Phenotype variable was then treated the same as the remaining phenotypes and run in the linear model with the remaining selected covariates.

Principal component analysis of the 17 selected phenotypes identified 6 principal components, explaining 85% of the variance across phenotypes. An adjusted study wide significance threshold, based on the conventional 5×10^−8^, was set at P≤8.33×10^−9^ (5×10^−8^ divided by 6 components). Additional consideration was given to the conventional genome wide threshold of P≤5×10^−8^ and a suggestive association threshold P≤1×10^−5^ for the purpose of comparison across phenotypes and for additional analyses.

To identify the peak SNPs for a genetic region, SNPs meeting at least the suggestive association threshold were trimmed based on Linkage Disequilibrium (LD) analysis. SNPs were sort ordered by p-value, and those in LD with R^2^ greater than 0.2 were considered to be in the same gene region for the purposes of identifying top SNPs for a genetic region.

While the dataset is highly AD specific, a separate analysis was performed with diagnosis as a covariate to distinguish SNP-phenotype associations not specific to AD (data not shown). Diagnosis was categorized as cognitively normal (CN), early mild cognitive impairment (EMCI), late mild cognitive impairment (LMCI), and AD.

### 2.5 Interaction Association with SNP and Diagnosis

The SNPxDX interaction analysis was performed through linear regression computed in R, with a phenotype as the dependent variable and SNP, diagnosis variable, and the interaction between the SNP and diagnosis variable as independent variables. The same covariates as before were applied (**Table 1**). Diagnosis was coded as an ordinal logistic variable, interpreted in four groups: CN, EMCI, LMCI, and AD. Ordinal coding interpreted these groups as having linear relationships between them with unknown spacing (CN < EMCI < LMCI < AD). The same thresholds and trimming methods as the main effect analysis above were applied on the interaction term of the model to determine significant interaction effects.

### 2.6 Functional Analysis

Variants were annotated for variant position and nearest gene using ANNOVAR (Version 2017-07-17). Study wide significant results were submitted to the Genotype-Tissue Expression (GTEx) Portal to determine *cis* expression quantitative trait locus (eQTL) effects. ADNI blood expression data was used to generate eQTL results within the dataset using the MatrixEQTL R package [16]. RegulomeDB [17] was referenced for potential regulatory effects for SNPs of interest.

## 3 RESULTS

### 3.1 Genome Wide Association Results

After LD trimming, a total of 27 genetic regions were identified as having study wide significance with at least one endophenotype. Figure 1 provides an overview of these top SNPs and their association with each endophenotype. The majority of findings are associated with the aggregate cognitive measures of Memory or Executive Function, with nine being study wide significant in other measures. Many associations showed effect directions suggesting a neuroprotective effect, with four findings apart from *APOE* showing a risk direction in association with the minor allele: rs2501374 and rs2506085 in an intergenic region on chromosome 1, rs9503939 intergenic on chromosome 19, and rs116622204 intronic to pseudogene *ZNF826P*.

**Figure 1:**
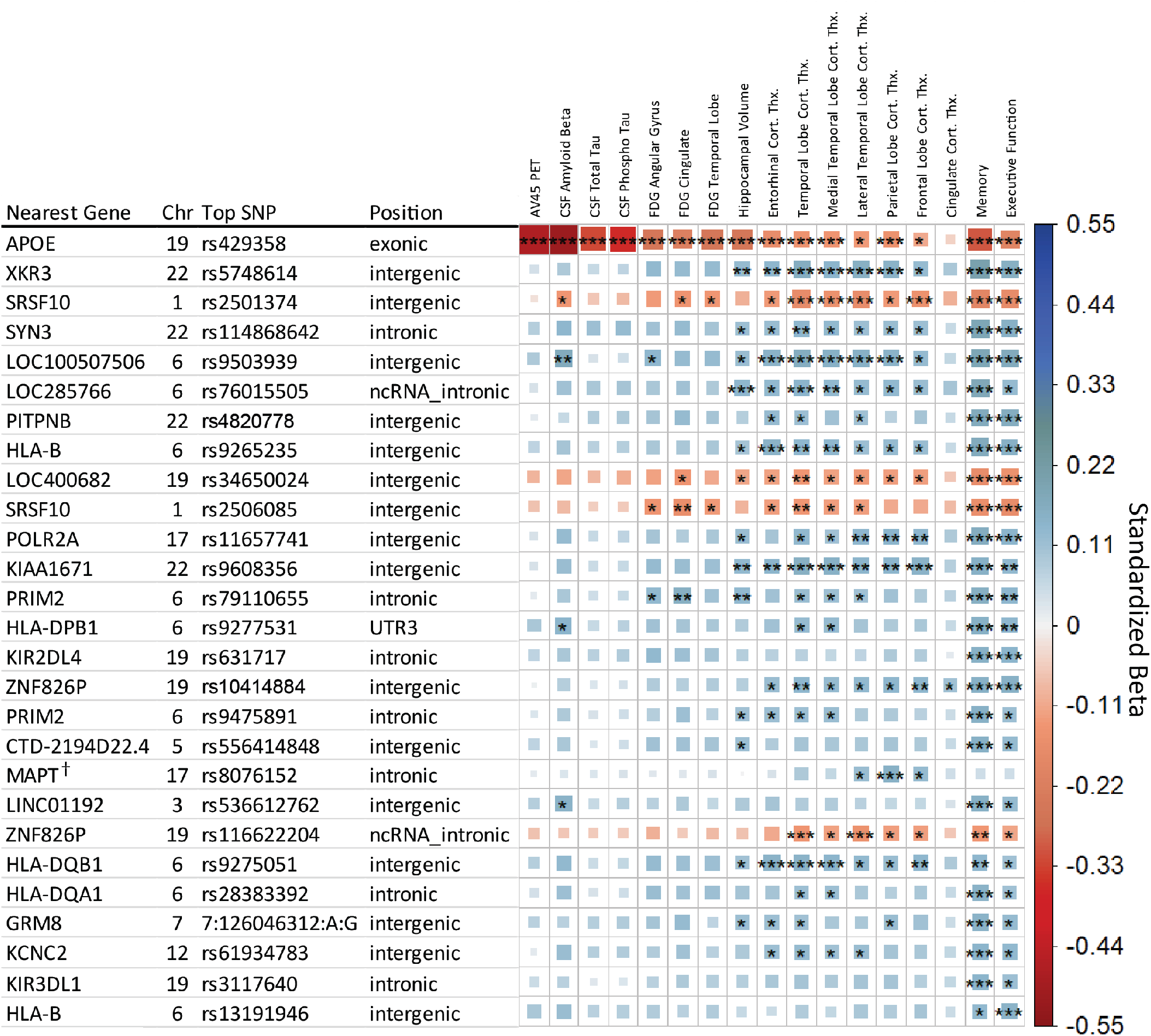
Matrix of main effect analysis results. Each row indicates the top SNP for a genetic region after LD trimming, and each column represents an AD endophenotype. Ordered based on minimum p-value across the row. The asterisks represent the P-value of the association with [***] indicating meeting the study wide significant threshold (P≤8.33×10^−9^), [**] the conventional genome wide threshold (P≤5×10^8^), and [*] the suggestive association threshold (P≤1×10^5^). The color and box size relates to the beta value effect size for a given association, with larger box size relating to distance from zero in either positive effect (blue, suggesting neuroprotective) or negative (red, suggesting neuropathological effect) direction. *†* Indicates the identified SNP retained significance with diagnosis as a covariate.

Including the *APOE* e2/e3/e4 allele as a covariate reduced the 27 study wide significant genetic regions to 16. Six SNPs maintained study wide significance in a non-cognitive measure, rs5748614 (near *XKR3*), rs2501374 (near *SRSF10*), rs9503939 (near LOC100507506), rs9608356 (near *KIAA1671*), rs8076152 (within *MAPT*), and rs116622204 (within *ZNF826P*). Including diagnosis as a covariate identified only one SNP that retained study wide significance, rs8076152, which is intronic to *MAPT* and associated with MRI measures of Parietal Lobe cortical thickness. A suggestive association was also found in Frontal and Lateral Temporal Lobe cortical thickness measures.

### 3.2 Known AD Associated Genetic Regions

Association analysis identified rs429358 (*APOE* e4 allele) on chromosome 19 as strongly associated with 13 of the 17 endophenotypes, with suggestive influence on two others. *APOE* e4 was most strongly associated with measures of amyloid-β, followed by measures of tau, glucose metabolism, and overall cognitive memory score. Strong associations were observed for surrounding SNPs in LD with *APOE* e4 in and near *TOMM40, APOC1*, and *NECTIN2* genes on chromosome 19.

Associations were also observed for regions in or near *HLA-DQA1, HLA-DPA1, and HLA-DRB1*, which have been identified in large-scale AD case/control studies [18].

### 3.3 SNPxDiagnosis Results

The SNPxDX model identified three study wide significant interaction SNPs. The first two were primarily associated with Parietal Lobe Cortical Thickness, rs1065272 located in the 3’ untranslated region of *BACH2* on Chromosome 6 and rs35823862 intronic to *EP300* on Chromosome 22. The *BACH2* SNP also showed suggestive associations with the Frontal Lobe and Lateral Temporal Lobe cortical thickness measures. A study wide significant SNP was also associated with FDG PET in the cingulate and is located in an intergenic region on Chromosome 6 near *PACRG-AS1*. The three SNPs showed a neuroprotective effect in later stages of AD, as represented in **Fig. 3** with the rs1065272 SNP in *BACH2*.

**Figure 2:**
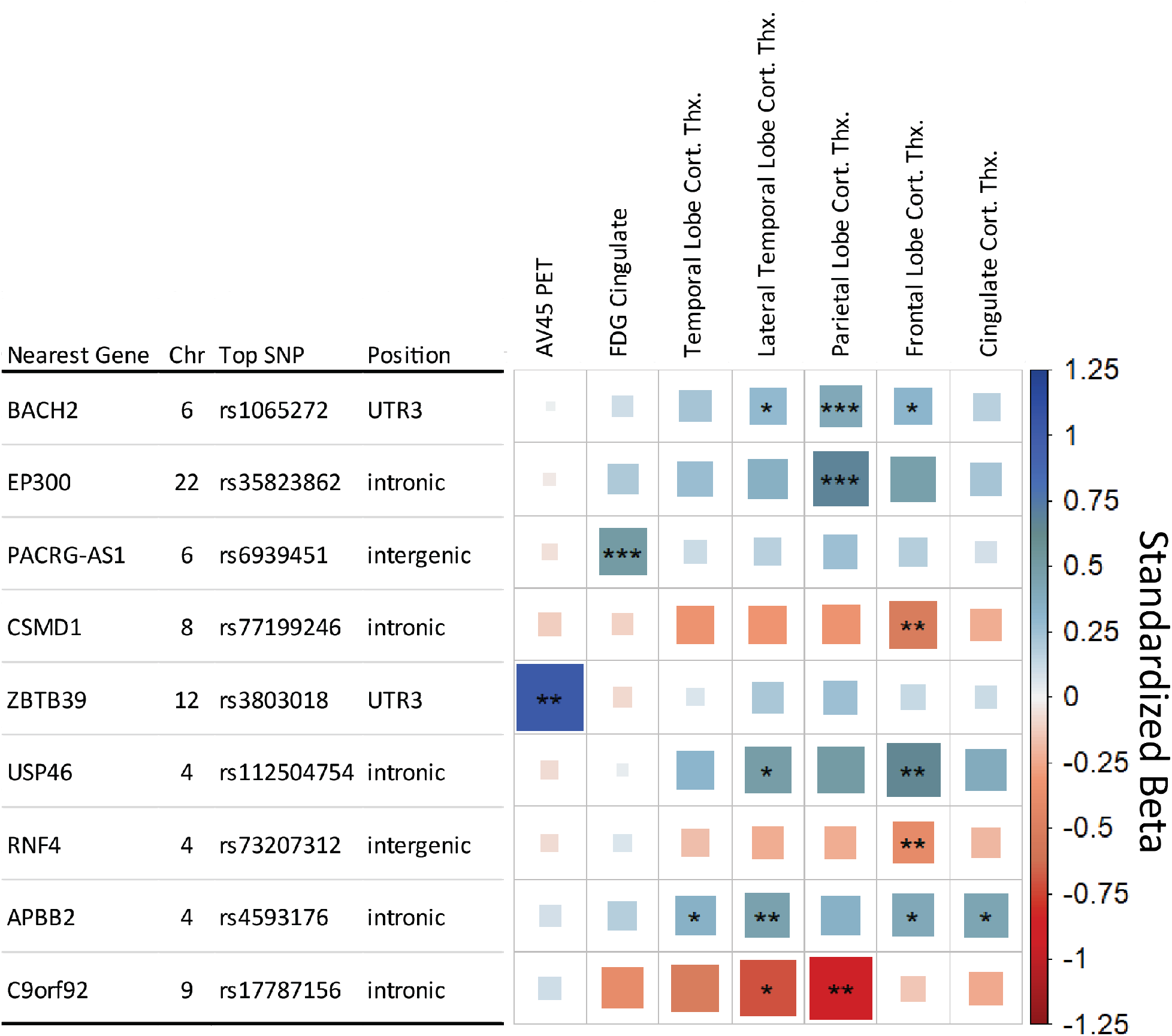
Matrix of SNP x Diagnosis analysis results. Each row indicates the top SNP for a genetic region after LD trimming, and each column represents an AD endophenotype. Endophenotypes showing no level of significance were removed for clarity. The asterisks represent the P-value of the association with [***] indicating meeting the study wide significant threshold (P≤8.33×10^−9^), [**] the conventional genome wide threshold (P≤5×10^8^), and [*] the suggestive threshold (P≤1×10^5^). The color and box size relates to the beta value effect size for a given association, with larger box size relating to distance from zero in either positive (blue, suggesting neuroprotective) or negative (red, suggesting neuropathological effect) direction.

**Figure 3:**
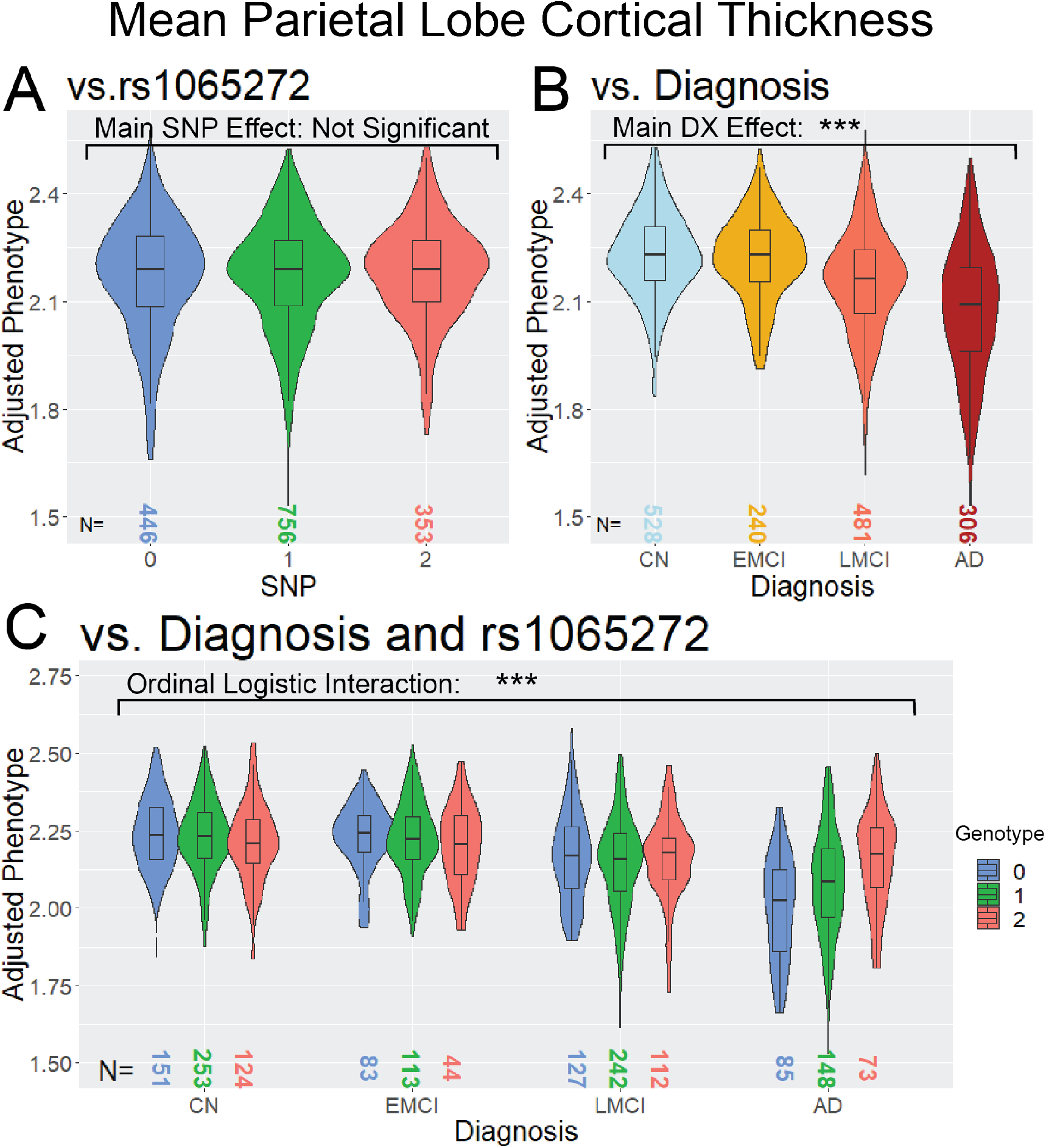
Violin and boxplot distribution of Parietal Lobe Cortical Thickness, stratified by (A) rs1065272 SNP, (B) Diagnosis, and (C) SNP and Diagnosis. (A) represents the Main Effect analysis, (B) the association with Diagnosis, and (C) the SNPxDX interaction association, with DX codes as an ordinal logistic variable of distinct categories with known ordinal relation (CN < EMCI < LMCI < AD). The asterisks represent the P-value of the association with [***] indicating meeting the study wide significant threshold (P≤8.33×10^−9^), [**] the conventional genome wide threshold (P≤5×10^8^), and [*] the suggestive threshold (P≤1×10^5^).

Additional six genetic regions met the less strict conventional genome wide significant threshold (P≤5×10^−8^) (Fig. 2). Including *APOE* genotype as a covariate in the SNPxDX model identified the same SNPs as without *APOE* adjustment.

## 3.4 Functional Results

The RegulomeDB analysis identified a score of 2b for the SNP rs2501374, indicating that there is evidence for transcription factor binding with a DNase footprint and DNase peak but no matched transcription factor. Other SNPs did not show suitable evidence in the RegulomeDB.

In the GTEx database and ADNI whole blood eQTL analysis, nine main effect SNPs were identified as being associated with altered expression. For the SNPxDX analysis, the SNP, rs3803018, was associated with altered *STAT6* expression levels in ADNI whole blood and altered *RDH16* expression levels in the Cerebellar Hemisphere in GTEx. **Table 2** provides a summary of eQTL findings between both approaches.

**Table 2:**
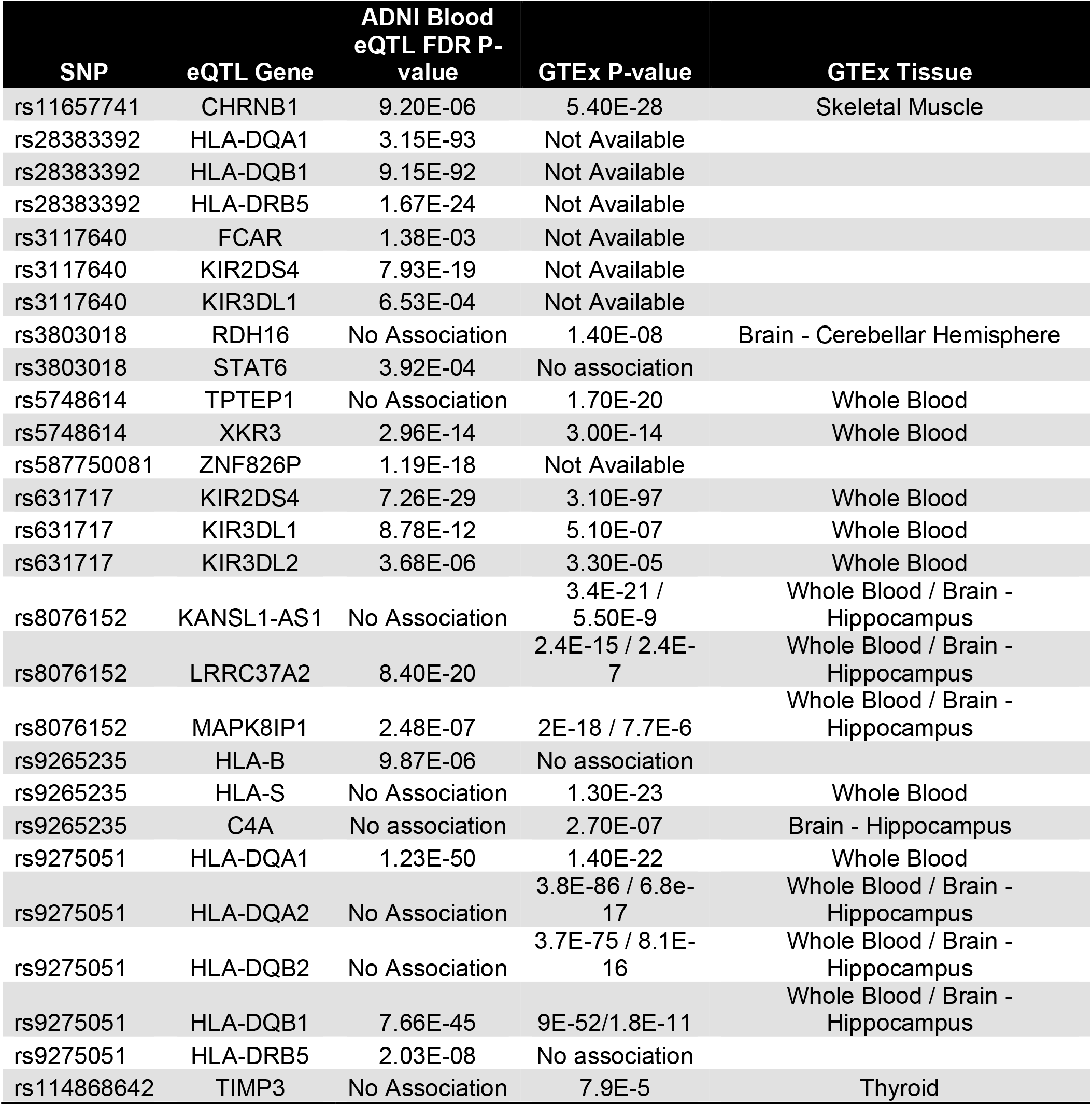
Summary of eQTL findings in ADNI whole blood analysis and GTEx database lookup.

## 4 DISCUSSION

Systematic analysis of genetic associations with key AD endophenotypes identified genetic regions previously associated with AD as well as several novel genetic associations. Analysis with a stage specific clinical syndrome approach further identified genetic effects that modulate endophenotypes in later stages of AD. The difference and variety of genetic associations in the two models highlights the need and value of more complex modeling for AD analysis. These variants are among many that have been identified in contributing to AD disease risk. Understanding how, where, and when these variants are affecting the disease through models such as those used in this study is key in developing an understanding of the disease and developing therapeutic approaches.

The identified SNPs identified in the main effect approach are largely in intergenic or uncharacterized regions, making functional analysis more challenging. The intergenic SNP rs2501374 proved particularly robust in this analysis. There is evidence of transcription factor binding in this region, though no direct eQTL evidence is available for this SNP. The nearest gene, *SRSF10*, is an alternative splicing gene which has general implications with AD pathogenesis [19]. *SRSF10* has known effects in enhanced lipogenesis [20] and affects alternative splicing of IL1RAP [21] which has been associated with AD pathology [22]. *XKR3* belongs to a family of phospholipid scramblases which have potential implications in apoptotic signaling [23], with eQTL evidence supporting a functional effect on expression in whole blood samples.

The HLA region has proven of interest in AD, particularly the HLA Class II region [18, 24-26] with evidence for the HLA Class I regions as well [27, 28]. Five independent HLA genetic regions were identified as having study wide significance in at least one endophenotype, with additional HLA Class I related markers in *KIR3DL1* and *KIR2DL4*. eQTL analysis identified association of these HLA region SNPs with HLA gene expression levels. However, study-wide significance is reduced in the HLA related SNPs when adjusting for *APOE* allele status, with only rs9265235 near *HLA-B* and rs9277531 near *HLA-DPB1* maintaining study wide significance in cognitive measures. Further study in how HLA related genes interact with AD pathology is warranted.

SNPs with study wide significance in a cognitive measure but suggestive association in other endophenotypes may provide insight as well. A SNP near *POL2RA* (rs11657741) shows a significant eQTL effect on *CHRNB1*, an acetylcholine receptor protein. There is evidence to suggest a role of the cholinergic system in AD [29]. *PRIM2* plays a role in DNA replication through Okazaki fragment formation, with DNA replication stress playing a potential role in AD pathology [30]. The SNP rs11486842 intronic to *SYN3* shows evidence of a small eQTL effect on *TIMP3*, which has been shown to regulate *APP* processing and *APOE* receptor proteolysis [31]. *SYN3* itself is involved in neurotransmitter release and synaptogenesis, and has been shown to be downregulated in hippocampal CA1 neurons in a tauopathy mouse model [32].

A SNP intronic to MAPT (rs8076152) is of potential interest, as it resides within the tau protein gene and is associated with neuroprotective effects in the parietal lobe. Evidence from this study suggests that the SNP is not directly related to AD as it retains its association with MRI measures when diagnosis is included as a covariate. While its position intronic to MAPT suggests it may have some effect on the tau protein gene, it may also influence various other genes, providing alternative functional outcomes.

The main effect analysis showed many significant associations with the composite cognitive measures. This is in part due to the cognitive scores having the highest N, resulting in better statistical detection power for association. MRI measures were close in sample size and corroborated many of the cognitive associations. Cognitive measures are not true endophenotypes but act as a surrogate quantitative measure for diagnosis, which may play a part in the scores representing the downstream effects of other phenotypic changes. Based on these results, utilizing a quantitative trait representative of diagnosis rather than a binary case/control could be of use in genetic association in AD. However, some caution is needed in interpreting the cognitive measures alone, as those measures may be confounded by non-AD factors.

Due to sample size differences, this dataset is best suited for evaluating MRI and cognitive phenotypes and lacks some power in PET and CSF measures for proper comparison across measures. However, the observed effect size can give insight into shared association as well as patterns of effect across endophenotypes. Despite sample size differences, SNPs rs2501374 and rs950939 showed suggestive association in CSF amyloid-β levels, consistent with study wide significance seen in those SNPs in MRI and cognitive measures.

Outside of *APOE* and its surrounding regions, there was very little significance seen in Amyloid-β and Tau biomarkers. This is in part due to the lower sample size of these measures, but other factors may contribute. Amyloid deposition in cognitively normal individuals [33] may affect detection power of AD specific genetic effects, and more complex models to account for these subjects may be needed. Additionally, polygenic risk score studies have provided evidence of common AD genetic markers having little contribution toward risk of amyloid deposition, distinct from *APOE* which contributes greatly toward amyloid deposition [34].

The analysis performed here for main effect does not well replicate the top genetic regions identified in large scale AD GWAS studies [18], outside of *APOE* and genes in the HLA class II region. The main difference is approach, with the large scale GWAS utilizing a case/control analysis where this study utilized an endophenotype approach. Despite the relatively smaller sample size, the endophenotype approach may provide more power for identifying genetic effects specific to AD related pathology. These genetic effects relate directly to AD mechanisms that contribute to differences in disease pathology, where case/control targets more generalized risk. In a highly heterogeneous disease like AD, a generalized approach may not have the power to identify genetic effects that will be important in a more personalized medicine approach.

The SNPxDX approach provides an alternative look at the genetic effects occurring in specific stages of AD, with study wide significant findings with AD specific implications. The top SNP in the *BACH2* gene region is located in its 3’ UTR, suggesting possible post-transcriptional regulatory effects of the SNP. *BACH2* is a transcriptional regulator involved in processes like NF-κB signaling, apoptosis in response to oxidative stress, nuclear import of actin, and CD4(+) T-cell differentiation. It has also been shown to be upregulated in β-amyloid-treated SH-SY5Y neuroblastoma cells [35]. The top genetic association in *EP300* does not have a clear functional effect, but is one of many strongly associated SNPs that blanket the *EP300* gene. *EP300* has been shown to be strongly associated as a candidate “Master Regulator” in AD genetic network [36].

SNPs that did not meet the study wide significant threshold in the SNPxDX results but met the less strict conventional genome wide significant threshold may still be of interest. A SNP intronic to the gene Amyloid Beta Precursor Protein Binding Family B Member 2 (*APBB2*), which has been previously associated with AD [37], was identified as being associated with a higher volume in the lateral temporal lobe in later AD stages.

As with many GWAS studies, these findings are limited by the selected population and sample size, and further replication in independent larger cohorts is warranted. A larger sample size would benefit both models, and with the SNPxDX approach, it would allow more sophisticated modeling of interactions in earlier stages of the disease that could not be interpreted with confidence given the current sample size limitations. Future studies will include gene and pathway enrichment, functional analysis in multi-omic datasets, and application of endophenotypic associations in a polygenic risk score model.

In summary, our findings show that an endophenotype approach can identify novel genetic associations with links to AD as well as provide insight into the identified associations. Endophenotypes allow for more complex models, such as the SNPxDX approach used here, which may be necessary in identifying genetic effects that contribute to AD risk and progression that might otherwise be missed in conventional models. The findings identified here may provide insight into potential therapeutic targets or provide insight into genetic risk. Expanding on the techniques utilized in this study through more comprehensive modeling and larger samples will likely provide further power for new discoveries.

## Data Availability

Data is available upon request to author.

## ACKNOWLEDGEMENTS

The support for this project was provided by the Alzheimer’s Disease Neuroimaging Initiative (National Institutes of Health Grant U01 AG024904) and ADNI DOD (Department of Defense award number W81XWH-12-2-0012). Additional support for data analysis was provided by NLM R01 LM012535, NIA R03 AG063250, NIA R01 AG19771, NIA P30 AG10133, NIA P30 AG072976, NIA R01 AG057739, NIA U01 AG024904, NLM R01 LM013463, R01 AG068193, T32 AG071444, U01 AG068057, NIGMS P50GM115318, NCATS UL1 TR001108, NIA K01 AG049050, NIA R01 AG061788, the Alzheimer’s Association, the Indiana Clinical and Translational Science Institute, and the IU Health-IU School of Medicine Strategic Neuroscience Research Initiative.

Data collection and sharing for this project was funded by the Alzheimer’s Disease Neuroimaging Initiative (ADNI) (National Institutes of Health Grant U01 AG024904) and DOD ADNI (Department of Defense award number W81XWH-12-2-0012). ADNI is funded by the National Institute on Aging, the National Institute of Biomedical Imaging and Bioengineering, and through generous contributions from the following: AbbVie, Alzheimer’s Association; Alzheimer’s Drug Discovery Foundation; Araclon Biotech; BioClinica, Inc.; Biogen; Bristol-Myers Squibb Company; CereSpir, Inc.; Cogstate; Eisai Inc.; Elan Pharmaceuticals, Inc.; Eli Lilly and Company; EuroImmun; F. Hoffmann-La Roche Ltd and its affiliated company Genentech, Inc.; Fujirebio; GE Healthcare; IXICO Ltd.;Janssen Alzheimer Immunotherapy Research & Development, LLC.; Johnson & Johnson Pharmaceutical Research & Development LLC.; Lumosity; Lundbeck; Merck & Co., Inc.;Meso Scale Diagnostics, LLC.; NeuroRx Research; Neurotrack Technologies; Novartis Pharmaceuticals Corporation; Pfizer Inc.; Piramal Imaging; Servier; Takeda Pharmaceutical Company; and Transition Therapeutics. The Canadian Institutes of Health Research is providing funds to support ADNI clinical sites in Canada. Private sector contributions are facilitated by the Foundation for the National Institutes of Health (www.fnih.org). The grantee organization is the Northern California Institute for Research and Education, and the study is coordinated by the Alzheimer’s Therapeutic Research Institute at the University of Southern California. ADNI data are disseminated by the Laboratory for Neuro Imaging at the University of Southern California.

The Genotype-Tissue Expression (GTEx) Project was supported by the [Common Fund](http://commonfund.nih.gov/GTEx/index) of the Office of the Director of the National Institutes of Health, and by NCI, NHGRI, NHLBI, NIDA, NIMH, and NINDS. The data used for the analyses described in this manuscript were obtained from the [GTEx Portal](http://www.gtexportal.org/) on 12/05/2019

